# A Machine Learning Framework to Detect Syncope using the Active Stand

**DOI:** 10.1101/2020.12.07.20245159

**Authors:** M. Carmody, C. Finucane, H. Nolan, C. O’Dwyer, M. Kwok, R. A. Kenny, C.W. Fan

**Affiliations:** Clinical Development, PQ Bypass, Inc., San Francisco, California, United States; Department of Medical Physics and Bioengineering, Mercer’s Institute for Successful Ageing, St. James’s Hospital, Dublin, Ireland; The Irish Longitudinal Study on Ageing (TILDA), Department of Medical Gerontology, Trinity College Dublin, Dublin, Ireland; Department of Gerontology, St. Vincent’s Hospital, Dublin, Ireland; Institute of Biomedical Engineering, University of New Brunswick, Fredericton, New Brunswick, Canada; Medicine for the Elderly, Mater Misericordiae, Dublin, Ireland

**Keywords:** predicting syncope, head-up tilt test, active stand test, pattern recognition

## Abstract

**Background:** Vasovagal syncope (VVS) is the most common form of syncope, accounting for 50-60% of unexplained syncope. Currently diagnosis is achieved via clinical assessment combined with the Head-Up Tilt Test (HUT).

**Aim:** To examine the utility of the active stand test (AS) to identify those with a positive HUT or diagnosis of VVS.

**Design:** Retrospective study of hemodynamic responses to AS.

**Methods:** Continuous blood pressure responses to AS from 101 patients attending a Falls and Blackouts Unit were acquired, including: 37 controls (CON), 64 with a clinical diagnosis of VVS (VVS+) (34 tilt-positive (HUT+) and 30 tilt-negative (HUT-)) with a mean age of 25 ± 9 years. A total of 33 hemodynamic features were extracted with a subset of these entered into linear discriminant classifier. Classification accuracy was assessed using N-fold cross-validation.

**Results:** Results indicated that it was possible to classify the outcome of the HUT with sensitivity of 58.8%, specificity of 63.3% and an accuracy of 60.9%. Using a multivariate classifier it was possible to identify those with a positive diagnosis of VVS with a sensitivity of 84.3%, specificity of 72.9% and an accuracy of 80.2%.

**Conclusion:** This study highlights the existence of a unique AS hemodynamic response characterised by autonomic hypersensitivity exhibited by young patients prone to VVS which is detectable using a multi-parameter machine learning framework. With further verification, this approach may have applications in syncope and falls diagnosis, population studies and the tracking of treatment efficacy.

## Introduction

Vasovagal syncope (VVS) is marked by a sudden decrease in blood pressure with an associated drop in heart rate, often resulting in syncope (faints) and falls in young and older people.^1^ It is a common symptom with up to half of the population suffering from at least one episode during their lifetime.^2^ Frequent reoccurrence of VVS can be severe enough to have a significant reduction in a patient’s quality of life, with a debilitating effect comparable to chronic arthritis.^3^

The standard diagnostic test for VVS is clinical assessment together with symptom reproduction via a positive Head-Up Tilt Test (HUT). HUTT is a resource intensive procedure requiring at least 40 minutes of testing and constant clinical supervision. HUT is typically performed in specialist referral centres and is therefore unsuitable for non-specialist application. In addition, large variations in test accuracy and reproducibility have been reported.^4,5^In the absence of a positive HUT, a clinical diagnosis of VVS is based on patient history.

When assessing a patient for unexplained syncope, the European Society of Cardiology recommends that a detailed history, physical examination, standard electrocardiogram (ECG) and supine and upright blood pressure (BP) measurements are performed.^6^ These BP measurements are commonly taken during the Active Stand Test (AS), a relatively simple procedure involving 5-10 minutes rest, followed by a further 3-5 minutes of standing. Hemodynamic variables, such as HR and BP, are recorded continuously throughout on a beat-to-beat basis.^7^

The ability to predict the outcome of the HUT would be of large clinical benefit, saving clinician time, patient discomfort and reducing costs^4^. Therefore, it is understandable that many previous attempts to identify biomarkers and predict HUT results can be found in the literature, ranging from patient history based methods to applying advanced signal processing and pattern recognition methods to hemodynamic responses.^4,8–10^

Heart rate (HR), along with blood pressure, is one of two main signals that were originally used to classify VVS. Using multivariate analysis,^8^ demonstrated that cardiovascular reactivity patterns during the first 10 minutes of baseline may be used to distinguish between syncope patients and controls with 93.3% sensitivity and 62.5% specificity. Similarly, some studies have demonstrated the degree of HR increase is greater in tilt-positive patients.^9^

In addition, the frequency spectrum of the RR interval tachogram has been analysed in an attempt to predict syncope. Kouakam *et al*. performed spectral analysis of low (LF) and high frequency (HF) bands, as well as the LF/HF power ratio during the last five minutes of the supine position and the first five minutes of head up tilt.^10^ In these patients, spectral analysis of HRV indicated that the LF/HF power ratio was highly correlated with HUT results after only five minutes of tilt with sensitivity and specificity both reaching 89%.

Furthermore, several researchers have used multi-feature predictors, including Virag *et al*. who demonstrated that the joint assessment of RR interval and SBP successfully predicted HUT outcome in 719 of 759 patients (sensitivity 95%, specificity 93%).^4^ However, this test required the implementation of the HUT and often included the prodromal symptom stage. Therefore, while it may reasonably predict the outcome of the HUT, it does not necessarily save much time or reduce patient discomfort.

Large variations in prediction accuracy and time are reported in the literature. However, it appears that no previous research has examined the predictive utility of the AS test in this regard.

The aim of this research is to examine the relationship between a patient’s AS response and the outcome of their HUT. We test the hypothesis that an alternative hemodynamic response to the AS test exists in those with a tendency towards VVS and that the outcome of the HUT can be predicted by analysing a patient’s hemodynamic response to the AS.

## Methods

This was a retrospective study using previously acquired data [11].

### Research Setting

All experimental procedures were conducted at the Fall’s and Blackout Unit (FABU) in St. James’s Hospital Dublin between 2008 and 2010. N = 64 patients with a history consistent with VVS (denoted VVS+), i.e. syncope precipitated by prolonged standing, fear, severe pain, emotional distress or instrumentation and associated with typical prodromal symptoms were recruited from FABU.

The cause of syncope was clinically evaluated and agreed upon by two independent, consultant geriatricians based on history, Sheldon scale and exclusion of other known causes of syncope in accordance with the European Society of Cardiology’s (ESC) guidelines. An additional 37 age-matched controls (CON) with no previous history of syncope were recruited from an existing database of healthy volunteers.

### Experimental Protocol

Initially, an AS test was conducted. This involved surface ECG and beat-to-beat blood pressure monitoring during 5 minutes of supine rest and 2-3 minutes standing. After the AS, syncope patients proceeded to a HUT test. Patients were secured safely on the HUT table and rested supine for an initial 5-minute stabilisation period. Following this, patients were tilted to 70° for 20 minutes. After this passive phase, 400µg of glycerine trinitrate (GTN) was administered sublingually, and the tilt was continued for a further 15 minutes (provocation phase). The test was terminated either at the end of the protocol or at the onset of syncope. Those with a positive test formed the HUT+ group, while those with a negative test formed the HUT-group. All investigations were performed in a quiet, temperature-controlled room (22-24°C).

### Equipment

Throughout both the AS and HUT test, the Finometer Pro (Finapres Medical Systems B. V., Netherlands) was used to monitor the hemodynamic activity of the patient. This system allows for the measurement of finger arterial pressure on a beat-to-beat basis, providing a non-invasive estimate of intra-arterial readings. It auto-corrects for the hydrostatic pressures associated with changes in hand position, as well as differences between brachial and finger-pressure waveform morphologies, by using a return-to-flow system. It has also passed both the AAMI/SP10 and BHS protocols for blood pressure measurement systems.^11^

In addition to this, a number of software packages were used to record, compile and analyse the data. These include BeatScope (Finapress Medical Systems B. V., the Netherlands), Excel (Microsoft, US-WA), MATLAB (Mathworks, US-MA) and SPSS (PASW Statistics 18, IBM, US-NY).

### Feature Extraction

A total of 33 features, as detailed in Appendix A Appendix A **Table 1** and Table 2, were extracted from the following signals: Systolic Blood Pressure (SBP), Heart Rate (HR), Stroke Volume (SV), Cardiac Output (CO) and Total Peripheral Resistance (TPR). Features were selected based on previous literature, visual inspection and control theory.

**Table 1:** Participant characteristics

|  | Units | HUT+<br>N = 34 | HUT-<br>N=30 | CON<br>N=37 |
| --- | --- | --- | --- | --- |
| Age | Years (Mean ± SD) | 25 ± 8.4 | 26 ± 7.0 | 24 ± 4.6 |
| Number Of Males | N (%) | 21 (62%) | 20 (67%) | 26 (70%) |
| Weight | kg (Mean ± SD) | 70 ± 14 | 70 ± 14 | 73 ± 15 |
| Height | cm (Mean ± SD) | 172 ± 11 | 169 ± 10 | 170 ± 10 |
| Number Of Faints | Median (Range) | 3.5 (2-72) | 6.5 (2-210) | NA |
| Duration Of Symptoms | Years (Mean ± SD) | 3.8 ± 5.0 | 3.3 ± 5.6 | NA |

**Table 2:** Significant features for HUT+ vs. CON

| Order of Significance | Feature | Units | HUT+<br>N = 34 | CON<br>N = 37 | P-Value |
| --- | --- | --- | --- | --- | --- |
| 1 | SV-B2SS | % (Mean ± SD) | -26.66 ± 11.54 | -16.14 ± 14.34 | 0.0022 |
| 2 | SBP-20s% | % (Mean ± SD) | 11.03 ± 16.31 | -0.09 ± 13.84 | 0.0023 |
| 3 | SBP-5s% | % (Mean ± SD) | -8.06 ± 13.86 | -17.54 ± 15.72 | 0.0115 |
| 4 | SBP-SSS | mmHg/sec | -0.07 ± 0.27 | 0.07 ± 0.31 | 0.0204 |
| 5 | HR-%I | % (Mean ± SD) | 57.49 ± 19.69 | 46.98 ± 23.47 | 0.0210 |
| 6 | HRV-SLF/SHF | Ratio (Mean ± SD) | 9.305 ± 13.80 | 14.560 ± 17.99 | 0.0309 |

Baseline features were established as the mean value from −60 to −30 seconds pre-stand. The final seconds pre-stand were not included in analysis due to significant signal noise present during the initial phase of the stand. The Steady-State values were taken as the mean value during 80 to 100 seconds post-stand.

Heart Rate Variability (HRV) features, based on frequency domain approaches, were calculated using Welch’s power spectral density (PSD) to estimate the PSD of both the Baseline and Steady-State stages. In addition to this, the root-mean-squared of successive differences (RMSSD) and standard deviation of successive differences (SDSD) of the pulse interval were calculated for each subject group. A list of calculation methods for each feature is available in Appendix A **Table 2**.

### Feature Selection

The Mann-Whitney U test was used to establish the significance of each feature between the subgroups HUT+ versus HUT- and VVS+ versus CON. A p-value of less than 0.05 was considered to be significant. Features were then eliminated based on their lesser significance when compared to the additional feature they were correlated with. A correlation coefficient of 0.6 or greater was used for the cut-off for elimination. The remaining, linearly independent features, were then ranked by the significance of their p-value. Based on these features, a linear discriminant function was formed, providing a distinction between the distribution of each groups’ features. This function was then used to classify additional patients based on their hemodynamic responses to standing and utilized to cross-validate and classify each patient into a specific group.

### Cross Validation and Classifier

The “k-fold cross-validation” classification technique was used to randomize the data set and divide it into k-partitions. A single partition was then selected to form the testing set, while the remaining k-1 partitions formed the training set. The classifier was trained and tested using this data. The process was then repeated, i.e. a new partition was selected from the k possibilities as the testing set and a new training set was then formed from the alternative k-1 partitions. This process was repeated k times, and the average results of the test sets formed an overall mean accuracy of the classifier.

In this study, a range of k-values were tested to classify the data. These were 2, 5 and 10. A shortcoming of k-fold cross validation is that each of the individual classifiers considered is trained on a subset of the data. Hence some information is ultimately wasted. With this in mind, an additional cross-validation method was also used in order to gain a range of accuracy for each classifier. Using this additional method, k was set to equal n, the number of patients in the dataset. This approach ensures that the maximum amount of training data (n-1) is available to each classifier. This method of n-fold cross-validation is commonly known as “leave-one-out cross-validation” (LOOCV) and has been proven to improve the performance of classifiers in many separate studies.^12–14^

Each of the features were entered into the linear classifier and the predictive powers of these univariate classifiers were examined. Following this, the features with the most predictive power were entered into a multivariate linear classifier in a stepwise fashion (highest predictive power first) to improve the overall accuracy.

### Assessing Classifier Performance

When applying cross-validation to a dataset with two distinct classes, there are four possible outcomes for each individual subject - true positive, false positive, true negative and false negative. For example, when classifying data from the Tilt-Positive (HUT+) and Control (CON) groups, the four possible outcomes are:

- **True Positive** (TP): A HUT+ subject classified as a HUT+ subject
- **False Positive** (FP): A CON subject classified HUT+ subject
- **True Negative** (TN): A CON subject classified as a CON subject
- **False Negative** (FN): A HUT+ subject classified as a CON subject

Classifier performance was based on the common statistical measures of sensitivity, specificity, positive predictive value, negative predictive value and overall accuracy. Where:

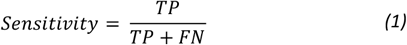

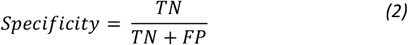

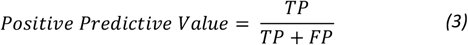

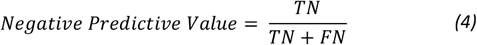

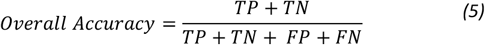

Finally, Receiver Operator Characteristic (ROC) curves were generated to document the clinical utility of the distinctions made with the classifiers developed. ROC curves plot sensitivity versus 1 – specificity over a range of prior probabilities (from 0 to 1) for the two classes and present a trade-off in costs between false positives and false negatives. This information can then be used to decide the threshold for different clinical requirements, e.g. screening versus pre-surgical diagnosis. The area enclosed by the ROC curve, or area under curve (AUC), has an important statistical property and is equivalent to the probability that the classifier will rank a randomly chosen positive instance higher than a randomly chosen negative instance.^14^

## Results

During testing, 34 of the VVS+ subjects were HUT+ and 30 were HUT-. This lead to the formation of three groups: HUT+, HUT- and CON. The participant characteristics collected from these individuals and their respective groups are detailed in Table III. As can be seen, the subjects included were a young cohort (mean age: 25 years) with a male predominance (66%). All features listed in Appendix Appendix A **Table 1** and Table 2 were analysed with respect to their ability to distinguish between each group. Tables IV, V and VI document each of the features that were found to have predictive utility for each group analysis (features with a p-value >0.05 are omitted).

**Table 3:** Significant features for HUT- vs. CON

| Order of Significance | Feature | Units | HUT-<br>N = 30 | CON<br>N = 37 | P-Value |
| --- | --- | --- | --- | --- | --- |
| 1 | SBP-20s% | % (Mean ± SD) | 12.29 ± 18.77 | -0.09 ± 13.84 | 0.0018 |
| 2 | HR-%I | % (Mean ± SD) | 61.55 ± 19.93 | 46.98 ± 23.47 | 0.0041 |
| 3 | SBP-5s% | % (Mean ± SD) | -5.30 ± 21.51 | -17.54 ± 15.72 | 0.0045 |
| 4 | SBP-10s% | % (Mean ± SD) | -22.57 ± 13.79 | -30.70 ± 15.44 | 0.0163 |
| 5 | HRV-SLF/SHF | Ratio (Mean ± SD) | 7.804 ± 7.798 | 14.560 ± 17.99 | 0.0261 |
| 6 | SBP-60s% | % (Mean ± SD) | 7.80 ± 10.16 | 2.08 ± 9.31 | 0.0336 |
| 7 | TPR-%D | % (Mean ± SD) | -40.03 ± 11.59 | -46.44 ± 12.81 | 0.0499 |

**Table 4:**
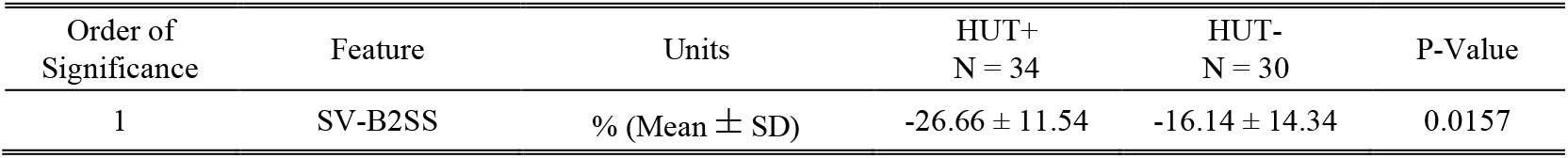
Significant features for HUT+ vs. HUT-

**Table 5:** HUT+ vs. HUT-: Optimum classifier

| N = 61 |  |  |  |  |  |  |  |
| --- | --- | --- | --- | --- | --- | --- | --- |
| Features | Sensitivity (%) | Specificity (%) | PPV (%) | NPV (%) | Accuracy (%) | Area Under Curve (%) | 95% Confidence Interval (%) |
| SV-B2SS | 58.8 | 63.3 | 64.5 | 57.6 | 60.9 | 67.6 | 54.5-80.8 |

Note: P/NPV = Positive/Negative Predictive Value

**Table 6:**
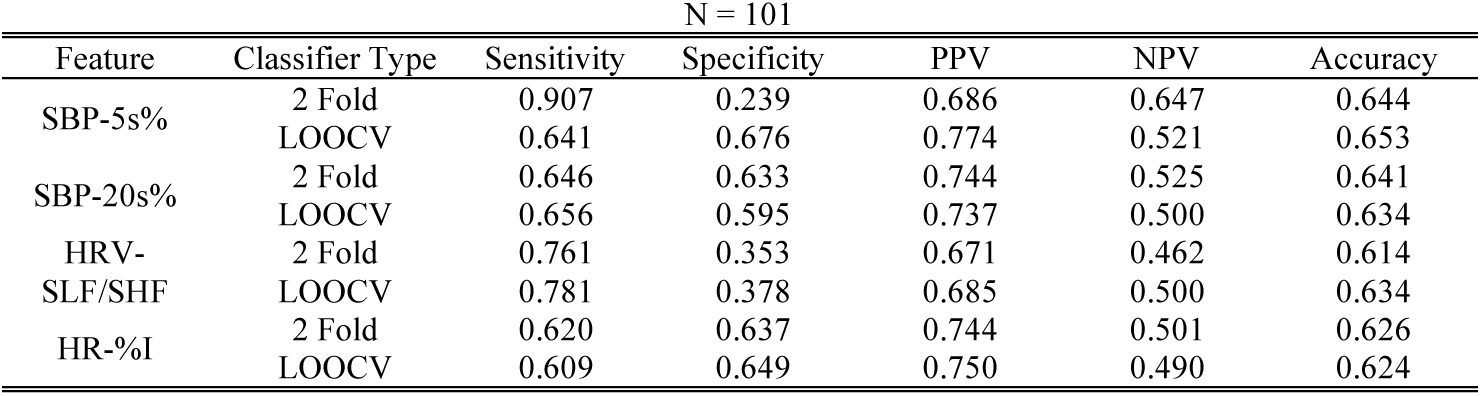
VVS+ vs. CON: Univariate classifier results

When comparing the HUT+ and HUT-groups, only one feature was found to have significant predictive power (p = 0.016). This was the percentage difference in SV from baseline to steady state (SV-B2SS). As can be seen in **Error! Reference source not found**., the mean SV response of the HUT+ group experiences a significantly larger (p < 0.05) reduction in stroke volume when compared with HUT-. When used in the univariate linear classifier, this feature performed poorly, producing a sensitivity of 58.8%, specificity of 63.3% and an accuracy of 60.9%, a relatively low predictive power (and Figure 5).

When analysing the VVS+ and CON groups, there were many significant features noted. The majority of which were functions of SBP, HR, SV and CO (Figures 1, respectively). As seen in the plots, VVS+ subjects experience a smaller drop in SBP after standing, followed by a greater overshoot and increased settling time. In addition, VVS+ subjects were also noted to have greater increases from baseline in both HR and CO. Finally, the VVS+ group experienced a greater reduction in SV from baseline to steady state.

**Figure 1:**
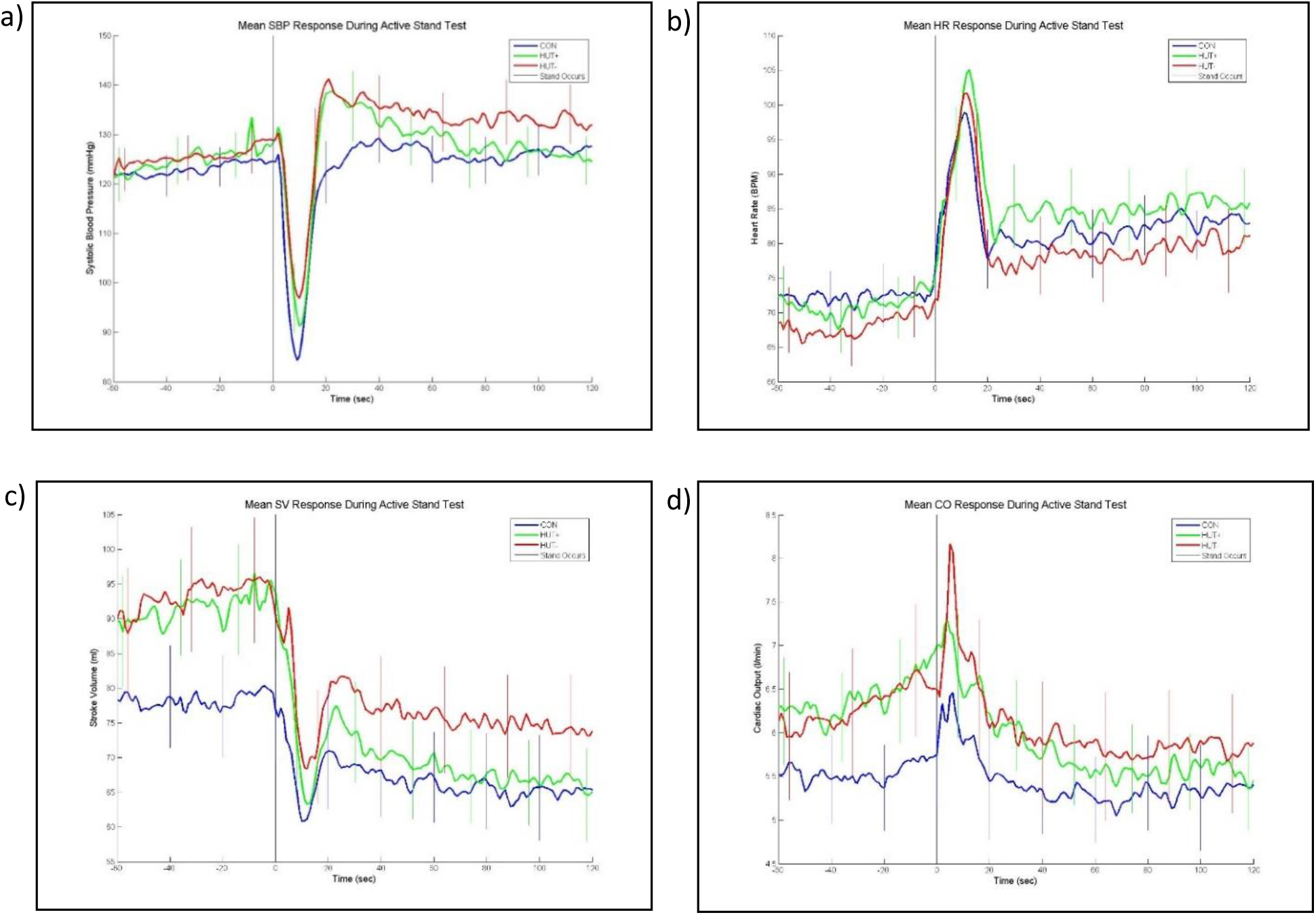
Mean response during active stand test including standard error for a) SBP b) HR c) SV d) CO

**Figure 2:**
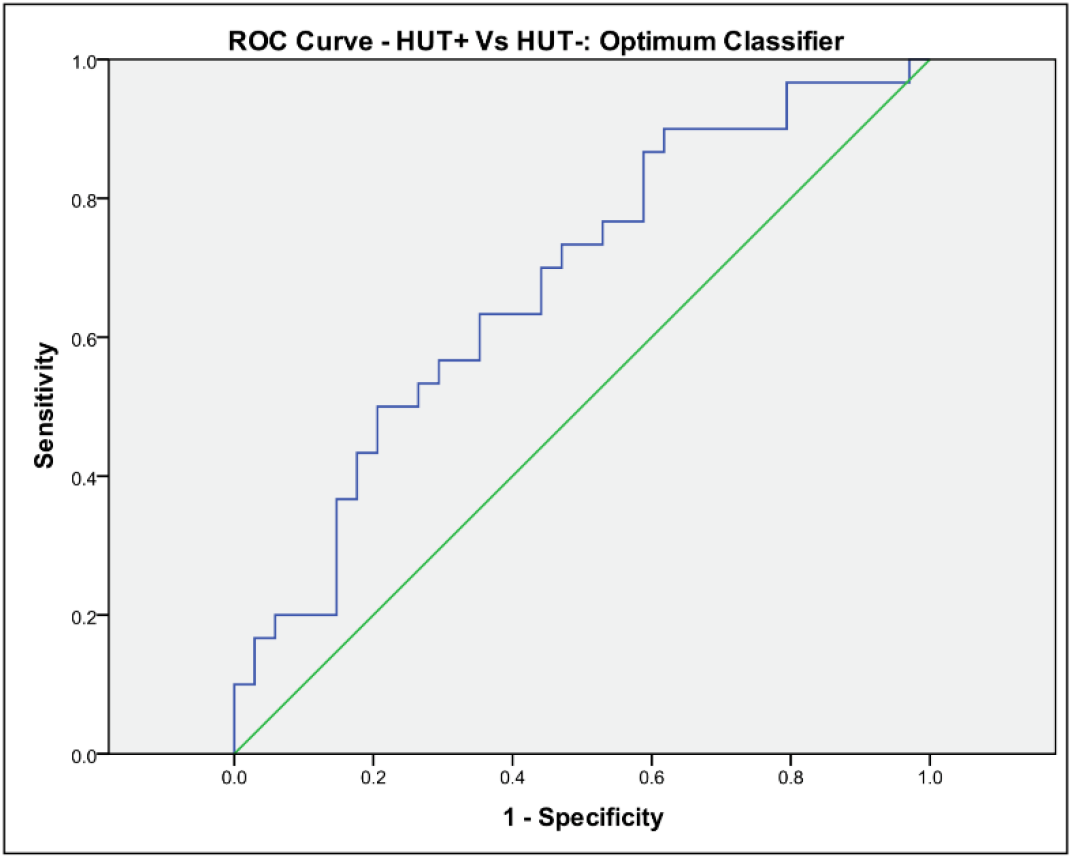
ROC curve for HUT+ vs. HUT-. Area under the curve is 67.6%.

**Figure 3:**
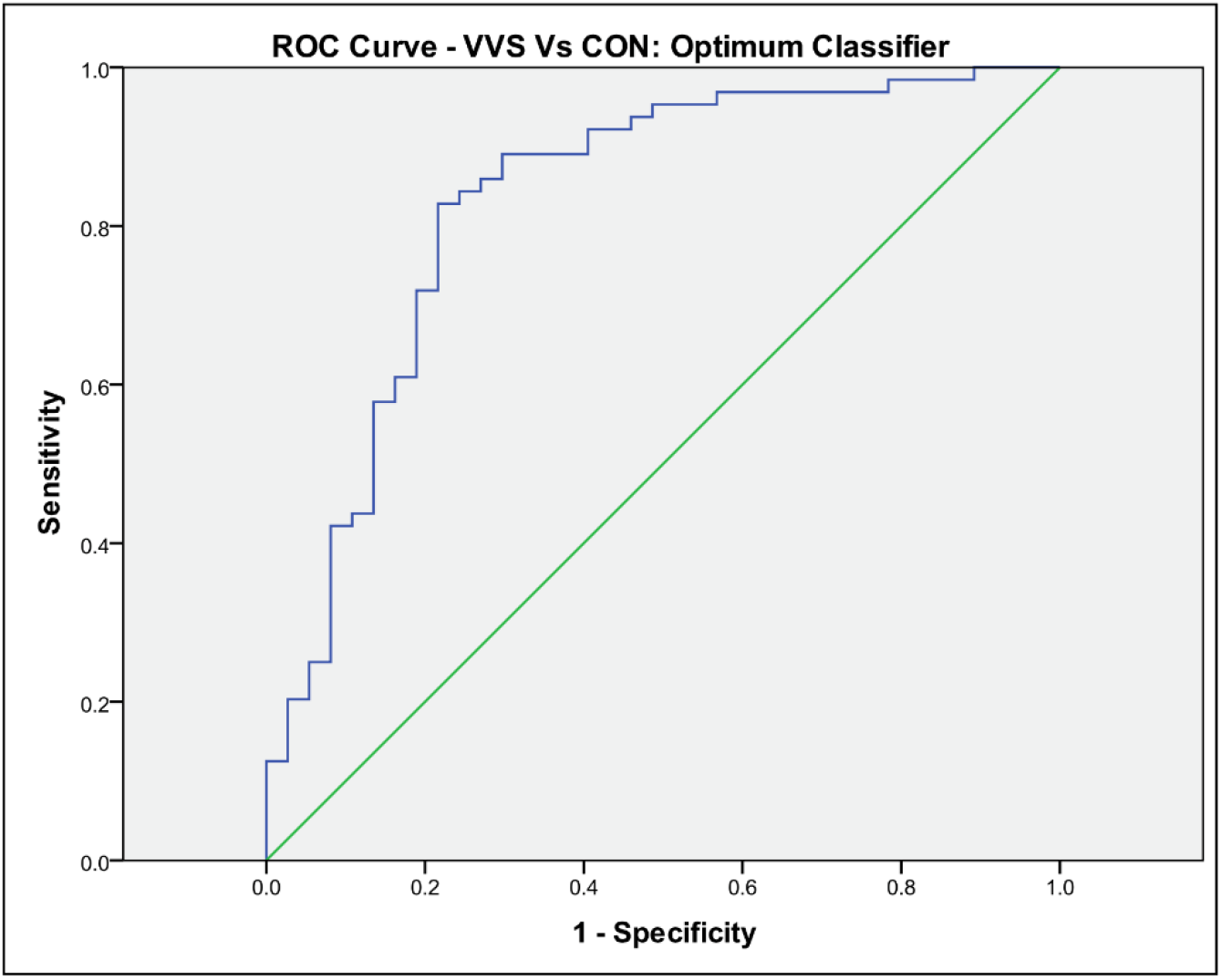
ROC curve for VVS+ vs. CON. Area under the curve is 83.2%.

Using the common features appearing in Tables IV and V, two univariate linear classifiers were developed to examine the predictive utility of each of the features with regards to the VVS+ and CON groups - a 2-fold classifier and a LOOCV classifier (Table VIII). A maximum accuracy of 65% was achieved using the percentage change in SBP from baseline at 5 seconds (SBP-5s%) when analysed using LOOCV.

**Table 7:**
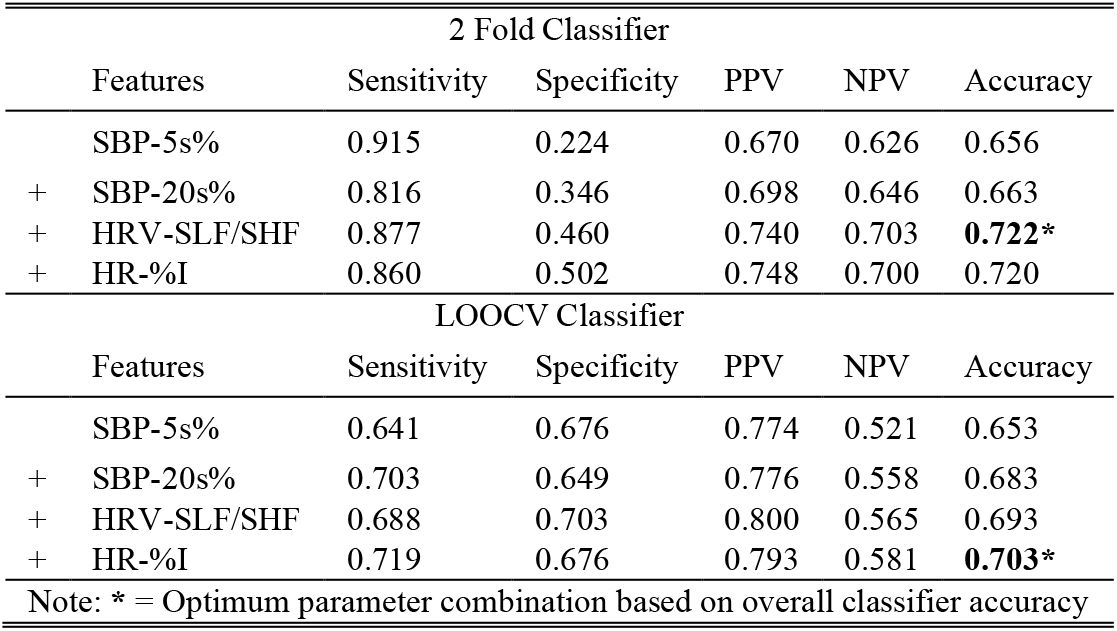
VVS+ vx. CON: Multivariate classifier results

| 2 Fold Classifier |  |  |  |  |  |  |
| --- | --- | --- | --- | --- | --- | --- |
|  | Features | Sensitivity | Specificity | PPV | NPV | Accuracy |
|  | SBP-5s% | 0.915 | 0.224 | 0.670 | 0.626 | 0.656 |
| + | SBP-20s% | 0.816 | 0.346 | 0.698 | 0.646 | 0.663 |
| + | HRV-SLF/SHF | 0.877 | 0.460 | 0.740 | 0.703 | **0.722\*** |
| + | HR-%I | 0.860 | 0.502 | 0.748 | 0.700 | 0.720 |
| LOOCV Classifier |  |  |  |  |  |  |
|  | Features | Sensitivity | Specificity | PPV | NPV | Accuracy |
|  | SBP-5s% | 0.641 | 0.676 | 0.774 | 0.521 | 0.653 |
| + | SBP-20s% | 0.703 | 0.649 | 0.776 | 0.558 | 0.683 |
| + | HRV-SLF/SHF | 0.688 | 0.703 | 0.800 | 0.565 | 0.693 |
| + | HR-%I | 0.719 | 0.676 | 0.793 | 0.581 | **0.703\*** |
| Note: * = Optimum parameter combination based on overall classifier accuracy |  |  |  |  |  |  |

Note: \* = Optimum parameter combination based on overall classifier accuracy

**Table 8:** VVS+ vs. CON: Optimum classifier

| N = 101 |  |  |  |  |  |  |  |
| --- | --- | --- | --- | --- | --- | --- | --- |
| Features | Sensitivity (%) | Specificity (%) | PPV (%) | NPV (%) | Accuracy (%) | Area Under Curve (%) | 95% Confidence Interval (%) |
| SBP-20s%;<br>HR-%I;<br>SV-B2SS;<br>SBP-5s%<br>CO-%I | 84.3 | 72.9 | 84.4 | 72.9 | 80.2 | 83.2 | 74.3-92.0 |

Note: P/NPV = Positive/Negative Predictive Value

Using the features above, two multivariate linear classifiers were also developed - a 2-fold classifier and a LOOCV classifier. The features with most predictive power were entered into a multivariate linear classifier in a stepwise fashion (highest predictive power first) based on the univariate analysis. The most accurate classifier (72.2%) was found using the 2-fold cross-validation method and uses only three features (Table IX).

A stepwise linear discriminant classification analysis using LOOCV was performed on each of the group relationships, and all of the previously established features were included. The stepwise method, as implemented in SPSS, resulted in an alternative classifier and an increase in accuracy from 72% to 80.2%.

The strongest combination of features contained the percentage change in SBP from baseline at 5 seconds and 20 seconds post-stand (SBP-5s%, SBP-20s%), the percentage increase from baseline of HR and CO (HR-%I, CO-%I) and the percentage difference in SV from baseline to steady state (SV-B2SS). This classifier produced a sensitivity of 84.3%, specificity of 72.9% and an accuracy of 80.2%. Table X and Figure 6 contain a summary of the results and the ROC curve for this classifier, respectively.

## Discussion

### Discussion of Findings

This study aimed to examine the utility of using cardiovascular responses to the AS to predict the outcome of the HUT using the patients’ hemodynamic response to the AS test. The most effective method established was a univariate classifier resulting in 58.8% sensitivity, 63.3% specificity and 60.9% accuracy.

However, when the HUT+ and HUT-subjects were merged to form a VVS+ group, a multivariate classifier was capable of distinguishing between this group and control subjects with 84.4% sensitivity, 72.9% specificity and 80.2% accuracy. This was the highest performing classifier and utilised five features: the percentage change in SBP from baseline at 5 seconds and 20 seconds post-stand (SBP-5s%, SBP-20s%), the percentage increase from baseline of HR and CO (HR-%I, CO-%I) and the percentage difference in SV from baseline to steady state (SV-B2SS).

### Comparison with Literature

Few studies have analysed the relationship between the AS response and the HUT outcome in adults. Using the AS, it has been found that the underlying cardiac autonomic mechanisms in vasovagal syncope may involve different autonomic patterns in subjects with a history of recurrent syncope.^15^ During active standing, heart rate and short-term complexity (alpha-1) increased in both groups (HUT+ and HUT-). In HUT+ patients, pNN50 decreased, whereas sympathovagal balance increased. The magnitude of change between positions of sympathovagal balance and alpha-1 was 6.1 and 4.8 times larger, respectively, in HUT+ than HUT-patients. However, the predictive power of these features was not reported.

Previous groups have attempted to predict the outcome of the HUT. Many researchers have developed multivariate classifiers capable of predicting syncope.^4,13,16^ Using the HUT, Ebden predicted syncope with a sensitivity of 93% and specificity of 88%.^13^ Similarly, it was found in Virag *et al* that syncope was predicted in a mean time of 128 ± 216s post-tilt with a sensitivity of 95% and specificity of 93%. However, prediction times varied from 0 to 30 minutes.^4^

Using the derivative of the ratio between RR interval and SBP (dRR/SBP), Mereu *et al*. was capable of predicting syncope with a sensitivity and specificity of 86.2% and 89.1% respectively.^16^ However, this was only possible within 44.1 ± 6.6s in advance of syncope.

An increase in HR has previously been observed by Mallat *et al*. in patients prone to VVS.^9^ They noted that an early sustained increase in HR ≤18 bpm during the first six minutes of tilt identified a negative test with 88.6% sensitivity and 100% specificity.

Additionally when comparing VVS+ subjects to CON, Pitzalis *et al*. noted that concurrent reductions of SBP during the first 15 minutes of tilt was indicative of a positive HUT with 93% sensitivity and 58% specificity.^17^ SBP was considered to be reduced when its beat-to-beat value was lower than the lowest baseline value. However, these observations could not be reproduced in this research due to the shorter duration of the AS.

It must also be noted that the tests in the literature were performed during the HUT and therefore spanned a much greater period of time, with some utilizing the entire length of the HUT.^4,13,16^ In addition to this, some studies administered vasodilatory drugs during testing.^18^ The use of these drugs during testing is likely to speed up the onset of syncope, and therefore it is not reasonable to compare it with a drug-free AS.

Finally, in clinical practice, a very wide range of HUT yields have been noted. Accuracy from 26% to 87% have been reported with the overall reproducibility of a negative response (85– 94%) being higher than that of an initial positive response (31–92%).^19^ When considering this, the extended time taken for these predictions and the fact the HUT apparatus is required, the classifier as described in this paper can be considered to be effective. Although it does not directly predict the outcome of the HUT on a specific day, it appears to distinguish between those prone to VVS and those who are not – a potentially more useful outcome, clinically.

### Pathophysiology

Given that the pathophysiology of VVS remains to be completely understood, one may only hypothesize the potential underlying mechanisms of these results. However, experimental data-to-date suggests that the VVS response is due to a sudden failure of the autonomic nervous system (ANS) to maintain adequate vascular tone and HR during orthostatic stress. This results in hypotension leading to cerebral hypoperfusion and loss of consciousness.^20^ While the contributing factors that lead to this ANS failure remain unclear, the subjects analysed within this study appear to exhibit the symptoms of a hypersensitive autonomic nervous system which over-respond to various stimuli, in this case, orthostatic stress.^21,22^ Furthermore, our results suggest that individuals prone to VVS have larger drops in SV on standing, suggestive of larger volume of blood pooling below heart level. This reaction may have some similarities with the Bezold-Jarisch reflex hypothesis. This theory states that fainting occurs due to cerebral hypoperfusion, as a result of profound vasodilation and bradycardia caused by blood pooling in the peripheral vasculature during prolonged standing.^23^ However, the question of whether this cardiac under-filling, coupled with excessive force of contraction, is sufficient to cause VVS has become a source of debate; several issues with this hypothesis have been highlighted.^13^

### Limitations

The subjects studied in this research were relatively young (mean age: 25 years). Given the heterogeneous nature of syncope sufferers, it is unlikely that this small, young cohort is representative of the entire syncope population. It is unknown whether or not the same hemodynamic pattern will apply to older patients prone to VVS, especially when considering the potential presence of comorbidities, such as OH, low BP and increased frailty. Therefore, these results must be considered within the context of the age range studied.

During the original research study in which this data was collected, the CON subjects were not subjected to the HUT. It is possible that a small proportion of this group may have experienced syncope.^24^

Finally, the AS is a very short test. Spanning only a few minutes, it may have limited the ability to establish a differentiating pattern. This may be notable in the lack of utility of the HRV features. These measurements are usually taken over a greater period of time and applying them to the AS may not reveal the true underlying variability.

## Conclusion

This paper examined the relationship between subjects’ hemodynamic response to standing, their clinical diagnosis of Vasovagal Syncope (VVS) and the outcome of a Head-Up Tilt Test (HUT). Here we have shown that while using the AS to predict HUT outcomes is limited, it is possible to distinguish between those with a clinical diagnosis of VVS and controls with 80.2% accuracy, a potentially more useful clinical outcome. This approach may have significant applications in population screening (e.g. population studies, military medicals) and tracking treatment efficacy. It also has the potential to be clinically applicable in situations where the HUT is not available or practical.

The existence of an alternative hemodynamic pattern exhibited in those with VVS during active standing may also provide some pathophysiological insight into the mechanisms of VVS. Here we note that those with VVS have evidence of increased levels of pooling and hypersensitive autonomic reflexes to standing.

## Data Availability

The confidential patient data is unavailable for access.

## Appendix

**Appendix A Table 9:**
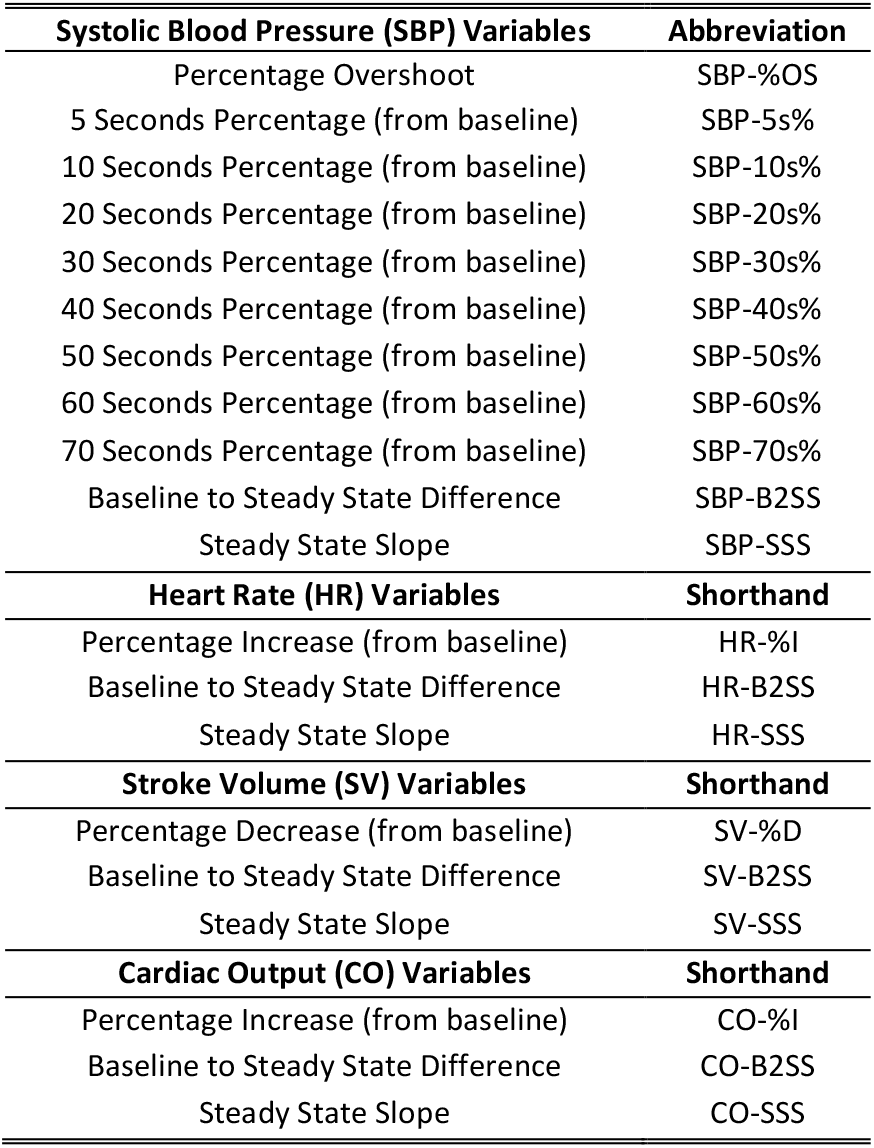
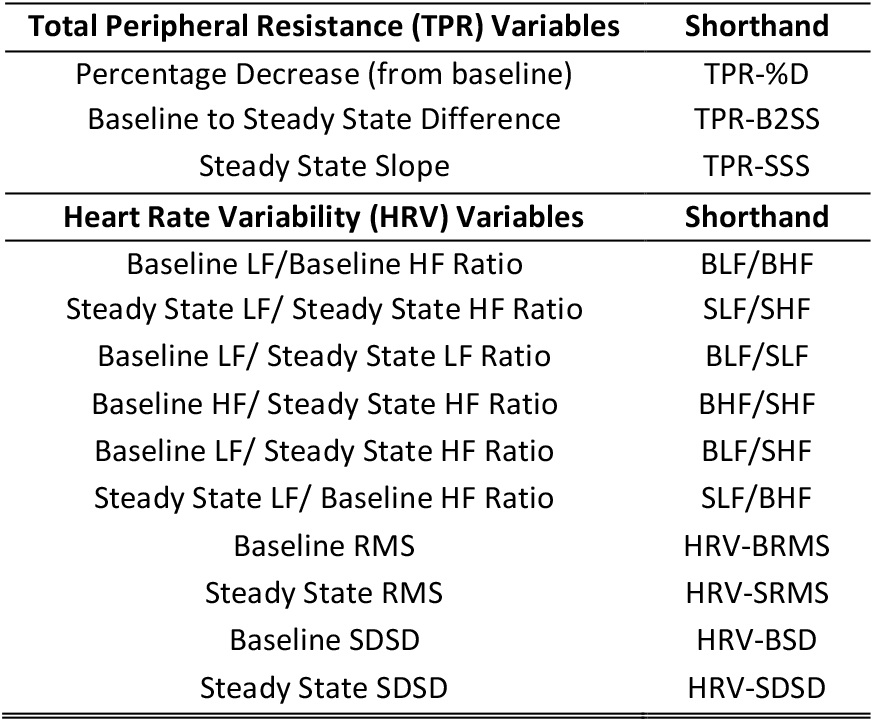
Features

**Appendix A Table 10:**
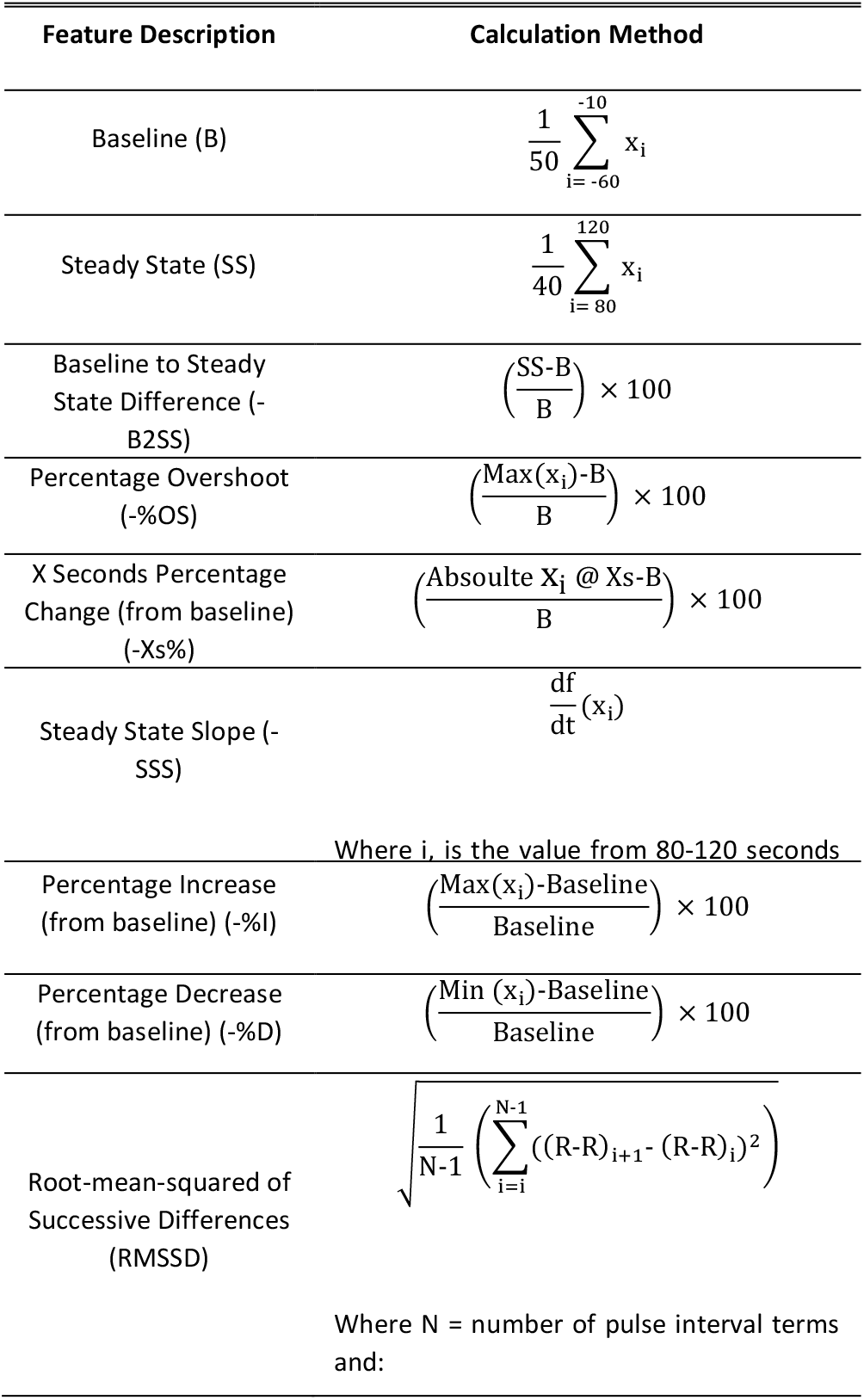
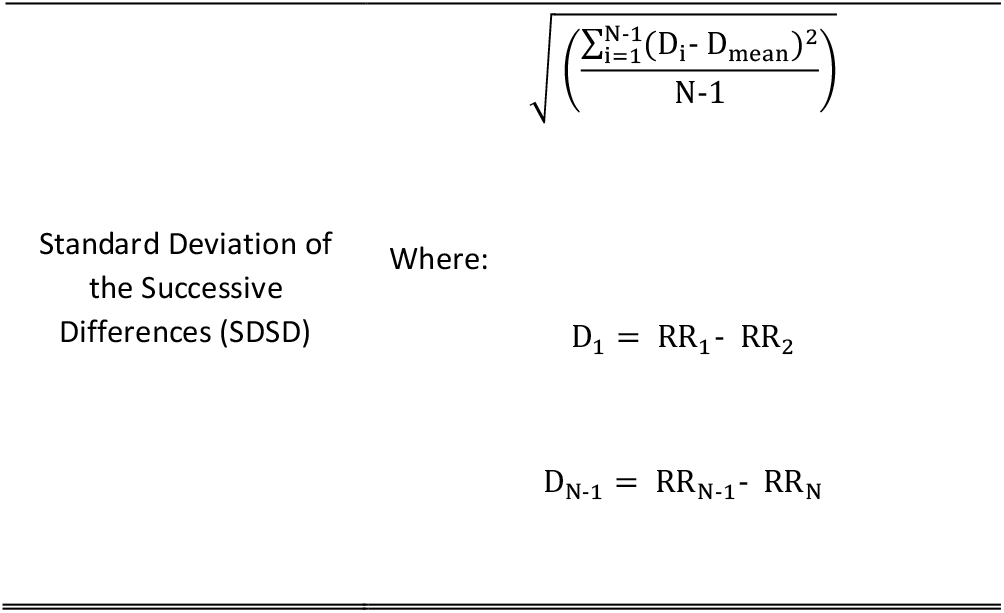
Feature calculations

## References

1. Brignole M. Diagnosis and treatment of syncope. Heart Br Card Soc. 2007;93:130–6.

2. Sutton R. Vasovagal syncope: prevalence and presentation. An algorithm of management in the aviation environment. Eur Heart J Suppl J Eur Soc Cardiol.1999;1 Suppl D:D109–113.

3. Linzer M, Yang EH, Estes NA 3rd, Wang P, Vorperian VR, Kapoor WN. Diagnosing syncope. Part 1: Value of history, physical examination, and electrocardiography. Clinical Efficacy Assessment Project of the American College of Physicians. Ann Intern Med. 1997;126:989–96.

4. Virag N, Sutton R, Vetter R, Markowitz T, Erickson M. Prediction of vasovagal syncope from heart rate and blood pressure trend and variability: experience in 1,155 patients. Heart Rhythm Off J Heart Rhythm Soc. 2007;4:1375–82.

5. Petkar S, Fitzpatrick A. Tilt-table testing: transient loss of consciousness discriminator or epiphenomenon? EP Eur. 2008;10:747–50.

6. Brignole M, Alboni P, Benditt D, Bergfeldt L, Blanc JJ, Bloch Thomsen PE, et al. Guidelines on management (diagnosis and treatment) of syncope. Eur Heart J. 2001;22:1256–306.

7. Task Force for the Diagnosis and Management of Syncope, European Society of Cardiology (ESC), European Heart Rhythm Association (EHRA), Heart Failure Association (HFA), Heart Rhythm Society (HRS), Moya A, et al. Guidelines for the diagnosis and management of syncope (version 2009). Eur Heart J. 2009;30:2631–71.

8. Naschitz JE, Rosner I, Rozenbaum M, Fields M, Isseroff H, Babich JP, et al. Patterns of cardiovascular reactivity in disease diagnosis. QJM Mon J Assoc Physicians. 2004;97:141–51.

9. Mallat Z, Vicaut E, Sangaré A, Verschueren J, Fontaine G, Frank R. Prediction of Head-Up Tilt Test Result by Analysis of Early Heart Rate Variations. Circulation. 1997;96:581–4.

10. Kouakam C, Lacroix D, Zghal N, Logier R, Klug D, Le Franc P, et al. Inadequate sympathovagal balance in response to orthostatism in patients with unexplained syncope and a positive head up tilt test. Heart Br Card Soc. 1999;82:312–8.

11. Wesseling KH. Finometer (TM) User’s Guide [Internet]. FMS, Finapres Medical Systerms BV; 2002. Available from: http://www.nuigalway.ie/psy/sub/manuals/finometer_1_ug.pdf

12. Bishop CM. Neural Networks for Pattern Recognition. New York, NY, USA: Oxford University Press, Inc., 1995.

13. Ebden M. Predicting Orthostatic Vasovagal Syncope with Signal Processing and Physiological Modelling. University of Oxford; 2006. 396 p.

14. Moran RJ, Reilly RB, Chazal P de, Lacy PD. Telephony-based voice pathology assessment using automated speech analysis. IEEE Trans Biomed Eng. 2006;53:468–77.

15. Vallejo M, Hermosillo AG, Infante O, Cárdenas M, Lerma C. Cardiac Autonomic Response to Active Standing in Adults With Vasovagal Syncope. J Clin Neurophysiol Off Publ Am Electroencephalogr Soc. 2015;32:434–9.

16. Mereu R, De Barbieri G, Perrone T, Mugellini A, Di Toro A, Bernardi L. Heart rate/blood pressure ratio as predictor of neuromediated syncope. Int J Cardiol. 2013;167:1170–5.

17. Pitzalis M, Massari F, Guida P, Iacoviello M, Mastropasqua F, Rizzon B, et al. Shortened Head-Up Tilting Test Guided by Systolic Pressure Reductions in Neurocardiogenic Syncope. Circulation. 2002;105:146–8.

18. Novak V, Novak P, Kus T, Nadeau R. Slow cardiovascular rhythms in tilt and syncope. J Clin Neurophysiol Off Publ Am Electroencephalogr Soc. 1995;12:64–71.

19. Fitzpatrick AP, Zaidi A. Tilt methodology in reflex syncope: emerging evidence. J Am Coll Cardiol. 2000;36:179–80.

20. Grubb Blair P. Neurocardiogenic Syncope and Related Disorders of Orthostatic Intolerance. Circulation. 2005;111:2997–3006.

21. van Dijk JG, Wieling W. Pathophysiological basis of syncope and neurological conditions that mimic syncope. Prog Cardiovasc Dis. 2013;55:345–56.

22. Grubb BP, Karas B. Clinical disorders of the autonomic nervous system associated with orthostatic intolerance: an overview of classification, clinical evaluation, and management. Pacing Clin Electrophysiol PACE. 1999;22:798–810.

23. Hainsworth R. Pathophysiology of syncope. Clin Auton Res Off J Clin Auton Res Soc. 2004;14 Suppl 1:18–24.

24. Furlan R, Piazza S, Dell’Orto S, Barbic F, Bianchi A, Mainardi L, et al. Cardiac Autonomic Patterns Preceding Occasional Vasovagal Reactions in Healthy Humans. Circulation. 1998;98:1756–61.

